# International expansion of a novel SARS-CoV-2 mutant

**DOI:** 10.1101/2020.03.15.20035204

**Authors:** Minjin Wang, Mengjiao Li, Ruotong Ren, Andreas Bråve, Sylvie van der Werf, En-Qiang Chen, Zhiyong Zong, Weimin Li, Binwu Ying

## Abstract

Letter to the editor. There is no abstract. The summary was showed: SARS-CoV-2 has inevitably mutated during its pandemic spread to cause unpredictable effects on COVID-19 and complicate epidemic control efforts. Here we report that a novel SARS-CoV-2 mutation (ORF3a) appears to be spreading worldwide, which deserves close attention.

## TO THE EDITOR

SARS-CoV-2 has inevitably mutated during its pandemic spread^1^ to cause unpredictable effects on COVID-19 and complicate epidemic control efforts. Here we report that a novel SARS-CoV-2 mutation appears to be spreading worldwide, which deserves close attention.

We detected 95 SARS-CoV-2 samples from Sichuan Province of China using next generation sequencing and acquired 13 whole genomes sequences, which were analyzed for sequence variation and evolution against 199 SARS-CoV-2 genomes publicly released in the GISAID EpiFlu™ database (https://www.gisaid.org/) and 7 genomes download from NGDC database (https://bigd.big.ac.cn/ncov). This study was approved by the Biomedical Research Ethics Committee of West China Hospital of Sichuan University (reference no. 193, 2020) and written informed consent was obtained from all patients.

Based on 10 high frequency mutations (mutant allele frequency >5%), these SARS-CoV-2 genomes can be classified into 5 main groups: original stain 1 and 4 variants with different mutations groups and clustering. The most common variants (Figure A, Group 1) exhibited both a missense mutation (ORF8:c.251tTa>tCa; present in 31.58% of the isolates) and a synonymous mutation (orf1ab:c.8517agC>agT; found in 30.62% of the isolates), suggesting a possible linkage between these two sites. Also, 3 subgroups were evolved in the main Group 1 by other 3 mutations. Group 2 was clustered together with 3 mutants including missense variant S: c.1841gAt>gGt, orf1ab upstream gene variant and synonymous_variant orf1ab: c.2772ttC>ttT. Group 3 viral isolates were much less frequent (11.48%) and characterized by a missense mutation (orf1ab:c.10818ttG>ttT). Group 4 viral isolates contained a novel missense mutation (ORF3a:c.752gGt>gTt) first found in a Chinese family. Notably, however, Group 4 viral isolates were most frequently found outside mainland China (23.28%; 27/116; *p*<0.01 by Fisher’s exact test). Additionally, Group 2 and Group 4 showed obvious aggregation in non-Chinese countries and regions.

**Figure.**
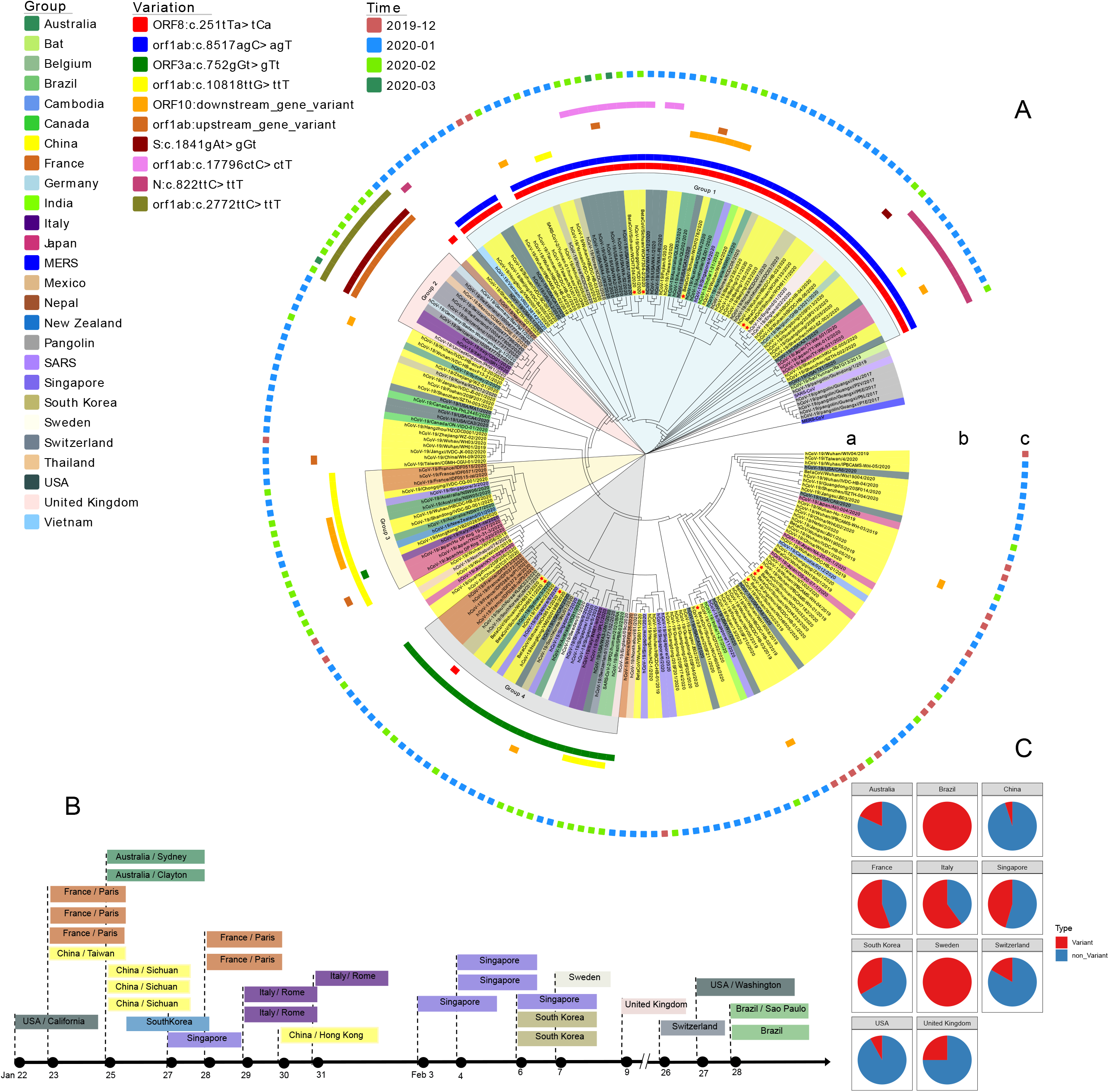
Maximum likelihood tree based on the whole genome sequences of 221 viral strains. A) 199 high quality genomes were collected from GISAID EpiFlu™ database, including 1 *Rhinolophus affinis* isolate, 6 *Manis javanica* isolates and 2 environmental isolates. 22 additional genomes were collected from other resource, including 7 genomes from NGDC (https://bigd.big.ac.cn/ncov), 13 genomes from WCH. SARS-CoV (NC_004718.3) and MERS-CoV (NC_019843.3) genomes sequence were downloaded from NCBI RefSeq database. MAFFT (version 7.543) was used for sequence alignment, and PhyML (version 3.0) was used to construct the evolutionary tree. Variation information of human SARS-CoV-2 genome was derived from NGDC. Mutations of 13 WCH genomes were analyzed using NGDC online tools (https://bigd.big.ac.cn/ncov/tool/variation-identify). B) Location and collection time of ORF3a:c.752gGt>gTt variant genomes. C) Composition of variant and non-variant genomes of ORF3a:c.752gGt>gTt in different countries.

The family in which the Group 4 variant was first observed in China (an older female and two young family members) returned to their hometown in Sichuan from Wuhan on January 20, 2020. By January 23, the mother had a fever and cough, and her two children developed these symptoms in the following days. Nucleic acid assays performed on their throat swabs tested positive for SARS-CoV-2 on January 25. None of these individuals traveled outside of China between the start of the COVID-19 epidemic and their return to Sichuan, but there the Group 4 variant first observed in this family has now demonstrated global dissemination.

We performed a timeline analysis using the sample collection dates reported in the GISAID EpiFlu™ database, and found that individuals infected with SARS-CoV-2 strain containing the Group 4 ORF3a mutant had reached the West Coast of the United States (Orange County, California) by January 22, 2020 at the latest. Immediately afterwards, and preceding or at nearly the same time as first Group 4 cases in Sichuan, additional isolates (Figure B) of this strain were reported in China (Taiwan), France (Paris), and Australia (Sydney and Clayton). According to the official records, these individuals either traveled from Wuhan, or traveled internationally prior to their disease onset. Group 4 ORF3a mutants were subsequently found in several other countries, including Singapore, South Korea, the United Kingdom and Italy. It should be noted that this mutant virus strain appears to be the most prevalent form of SARS-COV-2 in France, Italy,Brazil, and Singapore (Figure C).

Virus genome data from France indicate that SARS-CoV-2 strains carrying ORF3a:c.752gGt>gTt often have a S:c.1099Gtc>Ttc mutation in their S gene, which interacts with ACE2 to mediate viral entry into its host cells^3^, and is regarded as a critical factor for viral transmission and virulence^4, 5^. It is not clear if this mutation enhances host cell entry but this information would be of great importance in assessing the potential for increased virulence of Group 4 SARS-CoV-2 strains carrying this mutation. It is also not known how common this mutation is in Group 4 viral isolates from different geographical regions. Given the prevalence of Group 4 isolates in multiple countries, including France, Italy and South Korea, which is experiencing a rapidly growing epidemic, this information should be of significant interest.

At present, the SARS-CoV-2 epidemic in China seems to be diminishing in response to control efforts, but the rapid global spread of this new virus, and its mutants, has become a major health concern. Very little is known about how rapidly the SARS-CoV-2 genome mutates and how this affects transmission or disease severity. Better understanding of these factors should be useful in efforts to curtail the global and regional spread of this virus.

## Data Availability

The published genome data are abtained from the GISAID EpiFluTM database and NGDC database.

https://www.gisaid.org/

https://bigd.big.ac.cn/ncov

## Additional Contributions

We thank Dr Wan Xiong for polishing the manuscript language. We also thank Shuo Guo and Yanbing Zhou for collecting literature.

## Reference

1. www.who.int/emergencies/diseases/novel-coronavirus-2019/situation-reports/

2. Shu Y, McCauley J. GISAID: Global initiative on sharing all influenza data - from vision to reality[J]. Euro surveillance : bulletin Europeen sur les maladies transmissibles = European communicable disease bulletin. 2017,22(13). doi:org/10.2807/1560-7917.es.2017.22.13.30494

3. Letko M, Marzi A, Munster V. Functional assessment of cell entry and receptor usage for SARS-CoV-2 and other lineage B betacoronaviruses[J]. Nature microbiology. 2020. doi:org/10.1038/s41564-020-0688-y

4. Lu G, Wang Q, Gao GF. Bat-to-human: spike features determining ‘host jump’ of coronaviruses SARS-CoV, MERS-CoV, and beyond[J]. Trends in microbiology. 2015,23(8):468–478. doi:org/10.1016/j.tim.2015.06.003

5. Hamming I, Timens W, Bulthuis ML, Lely AT, Navis G, van Goor H. Tissue distribution of ACE2 protein, the functional receptor for SARS coronavirus. A first step in understanding SARS pathogenesis[J]. The Journal of pathology. 2004,203(2):631–637. doi:org/10.1002/path.1570

